# The causal role of low supervisor support in accidental blood exposure among hospital healthcare workers: a Directed Acyclic Graph (DAG) analysis of the STRIPPS cohort

**DOI:** 10.1101/2025.09.18.25336086

**Authors:** René Sosata Bun, Karim Aït Bouziad, Oumou Salama Daouda, Katiuska Miliani, Laura Temime, Mounia N Hocine, Pascal Astagneau

## Abstract

**Background:** Accidental blood exposures (AEB) are a major occupational hazard for healthcare workers (HCWs), with risk of bloodborne pathogen transmission. While organisational factors are known to influence safety, the specific causal pathways linking management quality to AEB risk remain poorly characterised.

**Objectives:** To investigate the causal pathways linking organisational factors, particularly supervisor support, to AEB risk among HCWs through stress and fatigue mediators using Directed Acyclic Graph (DAG) analysis.

**Methods:** Secondary analysis of the STRIPPS cohort study including 32 wards across four Paris university hospitals (n=730 HCWs). A DAG was constructed based on a literature review and previously published multivariate analyses to model causal relationships between organisational factors, psychological mediators, and AEB outcomes.

**Results:** AEB incidence was 4.1 per 100 visits overall, highest in intensive care units (7.1/100). DAG analysis showed that low supervisor support increased AEB risk through both direct and indirect pathways. Literature evidence indicated a protective effect of supervisor support on both stress and fatigue, while these psychological factors are strongly associated with increased AEB risk. Additional organisational factors including irregular work schedules, rotating shifts, and use of external personnel contributed to elevated AEB risk. Individual factors such as work overcommitment and presenteeism further amplified stress and fatigue pathways. The analysis explored multiple converging pathways from organisational and individual factors through psychological mediators to AEB risk.

**Conclusions:** Low supervisor support drives AEB risk through multiple interconnected pathways affecting stress and fatigue. Interventions targeting organisational support and psychological wellbeing could substantially reduce occupational injury risk among HCWs.

**Highlights:**

1. **What is already known about this subject?**
  - Accidental blood exposures (AEB) are a major occupational hazard for healthcare workers (HCWs), with significant risks of bloodborne pathogen transmission and psychological distress.
  - Organisational factors, including leadership and safety culture, are known to influence workplace safety, but their specific causal pathways to AEB remain poorly understood.
  - Stress and fatigue are recognised as mediators between work conditions and safety outcomes, but their roles in AEB have not been systematically modelled using causal methods.
2. **What are the new findings?**
  - Using a Directed Acyclic Graph (DAG) approach, we identified that low supervisor support increases AEB risk through both direct and indirect pathways mediated by stress and fatigue.
  - Fatigue showed a stronger association with AEB (OR 2.94–4.25) than stress (OR 1.12–1.53), highlighting its critical role in occupational safety.
  - Sickness presenteeism and work overcommitment were identified as key individual-level amplifiers of stress and fatigue, further increasing AEB risk.
  - Irregular work schedules and use of interim staff were organisational factors with substantial effects on AEB risk (RR > 3.0).
3. **How might this impact on policy or clinical practice in the foreseeable future?**
  - Healthcare organisations should prioritise supervisor training to improve supportiveness, which could reduce both psychological strain and AEB incidents.
  - Fatigue risk management systems and scheduling optimisations should be implemented to mitigate the strong effects of irregular shifts and long hours.
  - Policies discouraging presenteeism and promoting mental health support could break the cycle of fatigue and injury among HCWs.

## Introduction

Accidental blood exposures (AEB) represent a serious and prevalent occupational injury among healthcare workers (HCWs), representing a work-related hazard affecting an estimated 35-45% of global healthcare workforce annually[1]. These incidents carry substantial risks, including potential transmission of bloodborne pathogens such as HIV, hepatitis B, and hepatitis C, with global estimates suggesting that AEB account for 37% of hepatitis B, 39% of hepatitis C, and 4.4% of HIV infections among HCWs[1]. Beyond immediate infection risks, AEB generate significant psychological distress, costly post-exposure prophylaxis, and workforce disruption through medical leave and follow-up testing [2].

The healthcare environment is characterised by high workloads, time pressures, and emotional demands that create conditions leading to safety incidents[3]. Chronic exposure to workplace stressors emerges as psychological strain, particularly stress and fatigue, which may compromise adherence to safety protocols and increase injury risk[4]. Previous research has established associations between HCW stress and decreased compliance with safety measures, yet the specific mechanisms through which organisational factors translate into AEB risk remain not completely understood[5].

Organisational factors, particularly the quality of supervisory relationships, have emerged as critical determinants of workplace safety culture[6]. Supervisor support functions as a key resource that may temper job demands and protect against psychological strain. However, traditional analytical approaches often examine these relationships in isolation, failing to capture the complex interactions between organisational structures, psychological states, and safety outcomes.

Directed Acyclic Graphs (DAGs) provide a powerful framework for visualising and analysing causal relationships in complex systems, enabling identification of direct and indirect effects while accounting for potential confounding[7]. By employing DAG analysis, we can systematically map how organisational factors influence AEB risk through various mediating pathways, providing evidence-based targets for intervention.

This study aims to: (1) construct a comprehensive causal model linking organisational factors to AEB risk; (2) identify and quantify specific pathways through which supervisor support influences safety outcomes; (3) examine the mediating roles of stress and fatigue; and (4) provide actionable insights for organisational interventions to reduce occupational injury risk among HCWs.

## Methods

### Study Design and Data Source

This study represents a secondary analysis of data from the STRIPPS (*Stress at Work and Infectious Risk in Patients and Caregivers*) cohort, a prospective multicentre study conducted in four Paris university hospitals between February 2018 and July 2019. The study design and primary outcomes have been previously described**[8], [9]**.

The cohort included 32 hospital wards representing diverse specialties: medical (n=14), surgical (n=11), intensive care units (n=5), and obstetrics (n=2). Data were collected at baseline and at 4, 8, and 12 months through validated questionnaires assessing individual and organisational factors. Stress was measured using the Perceived Stress Scale (PSS-10)**[10]**, while fatigue was assessed using the Pichot scale**[11]**. Organisational factors were evaluated using adapted items from the Job Content Questionnaire**[12]**.

### Outcome Measures

AEB were defined as any percutaneous injury (needlestick, cut with sharp object) or mucocutaneous exposure to blood or other potentially infectious biological fluids, following CDC definitions**[13]**. Data were collected through self-reported incidents documented via standardised questionnaires at each time point. While self-reporting may underestimate true incidence, it captures events that HCWs perceive as significant, regardless of formal reporting**[14]**. Incidence rates were calculated as number of AEB per 100 ward visits during the study period.

### DAG Construction

We constructed our DAG through a systematic three-step process following recent methodological guidelines[15]:

#### Step 1: Foundation from STRIPPS Publications

We incorporated significant relationships from our previously published multivariate analyses examining: (a) determinants of stress and fatigue among HCWs[8] and (b) risk factors for AEB[9]. All factors with statistically significant adjusted risk ratios (p<0.05) were included as potential nodes.

#### Step 2: Literature Review

We conducted a rapid literature search to identify empirical relationships supporting our DAG construction:

- **Databases:** PubMed, Web of Science, Google Scholar
- **Time period:** January 2000 to January 2025
- **Search strategy:** Combined terms for (healthcare workers OR nurses OR hospital staff) AND (supervisor support OR managerial support OR organisational factors) AND (stress OR fatigue OR burnout) AND (needlestick OR blood exposure OR occupational injury)
- **Inclusion criteria:** Studies reporting multivariate-adjusted effect sizes in healthcare settings, using validated measurement instruments, published in peer-reviewed journals
- **Data extraction:** Adjusted effect sizes with confidence intervals, study characteristics, and confounders included in models

The search yielded 21 studies meeting inclusion criteria after full-text review. When multiple studies reported similar relationships, we prioritized longitudinal over cross-sectional designs and larger sample sizes.

##### Effect size transformations

When studies reported correlation coefficients (r) or t-statistics without corresponding odds ratios (OR) or risk ratios (RR), we calculated standardized effect sizes using established conversion formulas. For correlations, we applied Fisher’s z-transformation for confidence intervals. For t-statistics, we calculated Cohen’s d using sample sizes. When raw data were unavailable for transformation, effect sizes were reported as originally published and marked “NR” (not reported) for missing confidence intervals. All transformations are indicated with superscript “□” in tables.

#### Step 3: DAG Synthesis

The DAG was created using DAGitty version 3.1, following best practices for causal diagram construction[15]. We ensured:

- Absence of cycles (acyclic structure)
- Temporal ordering respected
- Inclusion of key confounders identified in literature
- Distinction between modifiable (organisational) and non-modifiable (individual) factors

Low supervisor support was selected as the primary exposure based on its consistent association with multiple outcomes and theoretical importance in occupational health frameworks. This DAG-based approach is particularly suited to occupational health research, where complex, multilevel determinants – from organisational policies to individual behaviours – interact to influence worker safety outcomes [15]

### Statistical Analysis

Descriptive statistics were calculated for all variables. AEB incidence rates were computed with 95% confidence intervals using Poisson distribution. The relationships depicted in our DAG were quantified using adjusted risk ratios from multivariate models accounting for ward type, time effects, and clustering by hospital[8], [9].

Effect sizes from the literature review were synthesised qualitatively rather than quantitatively due to heterogeneity in populations, measurements, and study designs. We report ranges of effects and identify consistent patterns across studies rather than pooled estimates.

All analyses were performed using R version 4.4.0 with packages including dagitty for DAG analysis and metafor for effect size transformations.

### Ethical Considerations

The STRIPPS study obtained both an agreement from the French Committee for the Protection of Persons (CPP) on 11/14/2017 and clearance from the French Data Protection Authority (CNIL) on 12/14/2017 (IDRCB N° 2017-A02939-44). This secondary analysis was covered under the original approval. All data were anonymised and no additional consent was required for this analysis.

## Results

### AEB Incidence Across Hospital Settings

During the 12-month study period, 108 AEB events were reported across 2,613 ward visits, yielding an overall incidence of 4.1 per 100 visits (95% CI: 3.4-5.0). Table 1 presents the distribution by specialty.

**Table 1.**
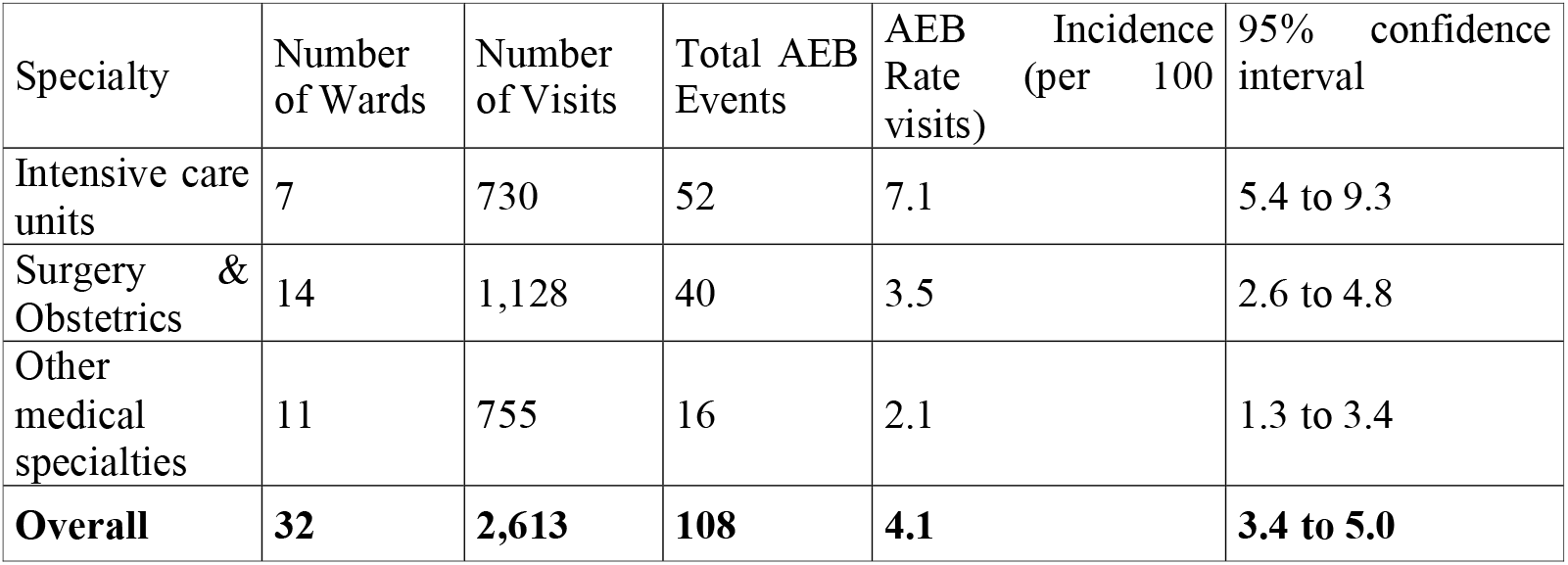
Distribution of AEB Events by Hospital Specialty in the STRIPPS cohort

| Specialty | Number of Wards | Number of Visits | Total AEB Events | AEB Rate (per 100 visits) | 95% confidence interval |
| --- | --- | --- | --- | --- | --- |
| Intensive care units | 7 | 730 | 52 | 7.1 | 5.4 to 9.3 |
| Surgery & Obstetrics | 14 | 1,128 | 40 | 3.5 | 2.6 to 4.8 |
| Other medical specialties | 11 | 755 | 16 | 2.1 | 1.3 to 3.4 |
| **Overall** | **32** | **2,613** | **108** | **4.1** | **3.4 to 5.0** |

**Figure 1.** presents the relationship between supervisor support levels and AEB incidence across the 32 participating wards. A negative correlation was observed between supervisor support scores and AEB rates, though this did not reach statistical significance (r = -0.246, p = 0.17).

**Figure 1.**
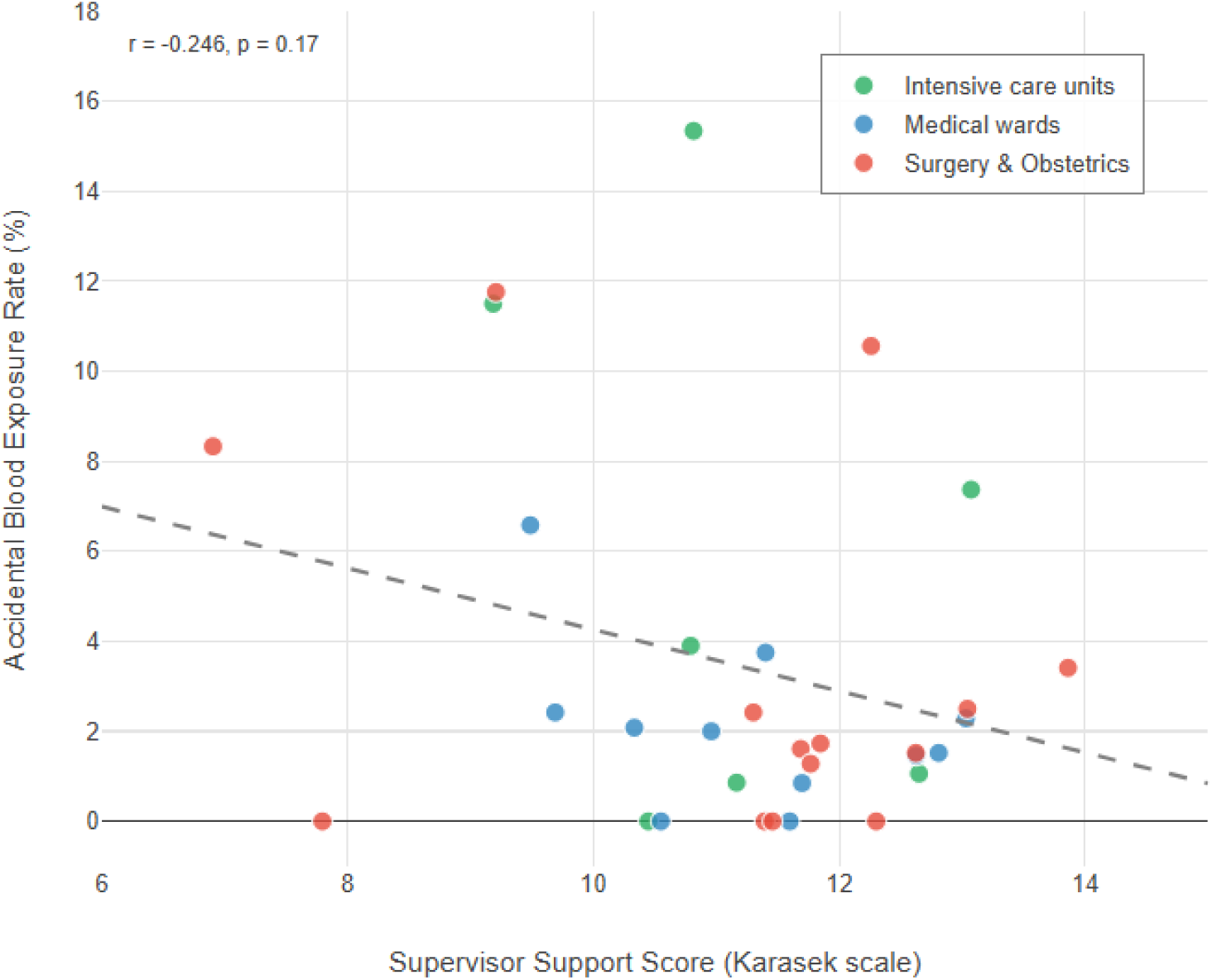
Supervisor Support and Accidental Blood Exposure Rates by Hospital Ward

### Literature Review: Quantifying Causal Pathways

Our systematic review identified 21 studies (total n=68,451 participants) from 12 countries/regions across 4 continents, examining relationships between organisational factors, psychological mediators, and AEB risk. Studies spanned from 2007 to 2024, using varied designs: cross-sectional (71%), longitudinal (14%), meta-analyses (10%) and case-crossover (5%),

**Table 2** summarises the key effect sizes identified in the literature for each causal relationship in our proposed model.

#### Supervisor Support as Protective Factor

Supervisor support demonstrated consistent protective effects across multiple outcomes:

- **Stress reduction:** β ranging from -0.23 to -0.26 across studies (Table 2A)[8], [16]
- **Fatigue reduction:** β = -0.34 in our cohort; β = -0.16 for low support in German nurses (Table 2B)[8], [21]
- **Direct AEB protection:** RR = 1.16 (95% CI: 1.06-1.28) for low support in STRIPPS[9]; correlation r = -0.17 in Chinese HCWs (Table 2 C)[28]

**Table 2A.** Literature review of effect sizes: occupational factors influencing stress, fatigue, and safety incidents among workers. Factors influencing perceived stress

| Causal Relationship | Proposed Mechanism | Source | Country | Design | n | Population | Effect Size | 95% CI | p-value |
| --- | --- | --- | --- | --- | --- | --- | --- | --- | --- |
| **Supervisor support**<br>(protective) | Social resources buffer job demands (**Org**) | Kiani, 2012[16] | Iran | Cross-sectional | n=189 | Company employees | $\beta = -0.26$ | NR | $p < 0.05$ |
| | | Daouda, 2022[8] | France | Longitudinal | n=695 | Hospital HCWs | $\beta = -0.23$ | -0.33 to -0.13 | $p < 0.001$ |
| **Coworker support**<br>(protective) | Peer assistance reduces burden (**Org**) | Tsai, 2012[17] | Taiwan | Cross-sectional | n=775 | Hospital staff | OR = 0.89 | 0.81 to 0.97 | $p < 0.05$ |
| | | Daouda, 2022[8] | France | Longitudinal | n=695 | Hospital HCWs | $\beta = -0.12$ | -0.24 to -0.01 | $p = 0.035$ |
| **Safety culture**<br>(protective) | Unsafe environment creates anxiety (**Org**) | Asefzadeh, 2017[18] | Iran | Cross-sectional | n=380 | Hospital nurses | $\gamma = -0.46$ | -0.73 to -0.19□ | $p < 0.001$ |
| | | Daouda, 2022[8] | France | Longitudinal | n=695 | Hospital HCWs | $\beta = 0.96$ | 0.07 to 1.84 | $p = 0.034$ |
| **Break cancellation** | Increased workload without recovery time (**Org**) | Daouda, 2022[8] | France | Longitudinal | n=695 | Hospital HCWs | $\beta = 1.78$ | 0.92 to 2.65 | $p < 0.001$ |
| **Work over-commitment** | Excessive investment depletes resources (**Ind**) | Hammig, 2012[19] | Switzerland | Cross-sectional | n=502 | Hospital employees | $\beta = 0.11$ | NR | $p < 0.05$ |
| | | Daouda, 2022[8] | France | Longitudinal | n=695 | Hospital HCWs | $\beta = 0.71$ | 0.63 to 0.78 | $p < 0.001$ |
| **Sickness presenteeism** | Working while ill increases strain (**Ind**) | Brborović, 2016[20] | Croatia | Cross-sectional | n=194 | Hospital staff | $d = 0.41$ | 0.21 to 0.61□ | $p = 0.015$ |
| | | Daouda, 2022[8] | France | Longitudinal | n=695 | Hospital HCWs | $\beta = 1.61$ | 0.55 to 2.66 | $p = 0.003$ |
| **Negative life events** | Cumulative life stressors overwhelm coping (**Ext**) | Daouda, 2022[8] | France | Longitudinal | n=695 | Hospital HCWs | $\beta = 2.27$ | 1.81 to 2.74 | $p < 0.001$ |

Effect sizes: *d*, Cohen's *d*; IRR, incidence rate ratio; NR, not reported; OR, odds ratio; RR, relative risk; *r*, correlation coefficient; $\beta$ , standardised regression coefficient; $\gamma$ , gamma coefficient. Level: Ind, individual; Org, organisational; Ext, external. □ Calculated/transformed from reported data.

**Table 2B.** Factors influencing fatigue

| **Causal Relationship** | **Proposed Mechanism** | **Source** | **Country** | **Design** | **n** | **Population** | **Effect Size** | **95% CI** | **p-value** |
| --- | --- | --- | --- | --- | --- | --- | --- | --- | --- |
| **Supervisor support**<br>(protective) | Social resources aid recovery<br>**(Org)** | Daouda, 2022[8] | France | Longitudinal | n=695 | Hospital HCWs | $\beta = -0.34$ | -0.45 to -0.24 | $p < 0.001$ |
| | | Petersen, 2023[21] | Germany | Cross-sectional | n=976 | Home-care nurses | $\beta = -0.159$ | -0.316 to -0.003 | $p < 0.046$ |
| **Perceived stress** | Chronic stress activation exhausts reserves **(Ind)** | Lang, 2022[22] | China | Cross-sectional | n=153 | Medical staff | $r = 0.61$ | 0.50 to 0.70 □ | $p < 0.001$ |
| | | Wang, 2025[23] | China | Cross-sectional | n=1,004 | Doctors and nurses | OR = 1.98 | 1.43 to 2.74 | $p = 0.001$ |
| **Sickness presenteeism** | Continued work despite illness depletes energy **(Ind)** | Daouda, 2022[8] | France | Longitudinal | n=695 | Hospital HCWs | $\beta = 4.03$ | 2.84 to 5.22 | $p < 0.001$ |
| | | Li, 2022[24] | China | Cross-sectional | n=2,968 | Nurses | $r = 0.201$ | 0.17 to 0.24 □ | $p < 0.001$ |
| **Work over-commitment** | Excessive investment exhausts reserves **(Ind)** | Aronsson, 2017[25] | Global | Meta-analysis | 25 studies (n=35,735) | Various groups | OR = 1.86 | 1.37 to 2.52 | $p < 0.05$ |
| | | Daouda, 2022[8] | France | Longitudinal | n=695 | Hospital HCWs | $\beta = 0.65$ | 0.04 to 1.22 | $p < 0.001$ |
| **Use of interim nurses** | Staff instability increases workload **(Org)** | Daouda, 2022[8] | France | Longitudinal | n=695 | Hospital HCWs | $\beta = 1.96$ | 0.35 to 3.56 | $p = 0.017$ |
| **Safety culture**<br>(protective) | Poor safety climate increases vigilance demands **(Org)** | Poursadeqiyan, 2020[26] | Iran | Cross-sectional | n=143 | Hospital nurses | $r = -0.29$ | -0.43 to -0.13 □ | $p < 0.05$ |
| | | Daouda, 2022[8] | France | Longitudinal | n=695 | Hospital HCWs | $\beta = 1.76 \square$ | 0.81 to 2.70 | $p < 0.001$ |
| **Female gender** | Gender-specific physiological and social factors **(Ind)** | Poursadeqiyan, 2020[26] | Iran | Cross-sectional | n=143 | Hospital nurses | $d = 0.50 \square$ | 0.26 to 0.74 □ | $p = 0.004$ |
| | | Daouda, 2022[8] | France | Longitudinal | n=695 | Hospital HCWs | $\beta = 1.52$ | 0.40 to 2.63 | $p = 0.008$ |
| **Longer commuting time** | Extended travel reduces recovery time **(Ext)** | Daouda, 2022[8] | France | Longitudinal | n=695 | Hospital HCWs | $\beta = 1.82$ | 0.80 to 2.83 | $p < 0.001$ |
| | | Mohd Suadi Nata, 2024[27] | Malaysia | Cross-sectional | n=212 | Full-time workers | $r = 0.42$ | 0.16 to 0.63 □ | $p < 0.05$ |

Effect sizes: *d*, Cohen's *d*; IRR, incidence rate ratio; NR, not reported; OR, odds ratio; RR, relative risk; *r*, correlation coefficient; $\beta$ , standardised regression coefficient; $\gamma$ , gamma coefficient. Level: Ind, individual; Org, organisational; Ext, external. □ Calculated/transformed from reported data; □ 'Not agree' reference category.

**Table 2C.** Factors influencing accidental exposure to blood (AEB)

| **Causal Relationship** | **Proposed Mechanism** | **Source** | **Country** | **Design** | **n** | **Population** | **Effect Size** | **95% CI** | **p-value** |
| --- | --- | --- | --- | --- | --- | --- | --- | --- | --- |
| **Supervisor support** | Poor support reduces safety behaviour (**Org**) | Wang, 2019[28] | China | Cross-sectional | n=1,956 | Hospital HCWs | r = -0.17 | -0.21 to -0.13□ | p < 0.01 |
|  |  | Bun, 2024[9] | France | Longitudinal | n=730 | Hospital HCWs | RR = 1.16 | 1.06 to 1.28 | p < 0.001 |
| **Irregular work schedule** | Schedule unpredictability disrupts routines (**Org**) | Trinkoff, 2007[29] | USA | Longitudinal | n=2,273 | Registered nurses | OR = 1.46 | 1.15 to 1.86 | p < 0.01 |
|  |  | Bun, 2024[9] | France | Longitudinal | n=730 | Hospital HCWs | RR = 3.18 | 1.83 to 5.11 | p < 0.001 |
| **Rotating shifts** | Circadian disruption impairs alertness (**Org**) | Hassanipour, 2021[30] | Global | Meta-analysis | 43 articles (n=14,817) | Healthcare personnel | OR = 2.16 | 1.47 to 3.15 | p < 0.001 |
|  |  | Bun, 2024[9] | France | Longitudinal | n=730 | Hospital HCWs | RR = 3.11 | 1.72 to 4.83 | p < 0.001 |
| **Use of external personnel** | Staff unfamiliarity with procedures (**Org**) | Bun, 2024[9] | France | Longitudinal | n=730 | Hospital HCWs | RR = 2.02 | 1.19 to 3.35 | p = 0.010 |
| **Perceived stress** | Cognitive overload impairs attention (**Ind**) | Wang, 2019[28] | China | Cross-sectional | n=1,956 | Hospital HCWs | r = 0.53 | 0.50 to 0.56□ | p < 0.01 |
|  |  | Amores-Lizcano, 2024[5] | Spain | Cross-sectional | n=475 | Healthcare workers | OR = 1.12□ | 1.05 to 1.19 | p < 0.001 |
| **Fatigue** | Reduced alertness and motor coordination (**Ind**) | Fisman, 2007[31] | USA/Canada | Case-crossover | n=350 | Medical trainees and HCWs | IRR = 2.94 | 1.71 to 5.07 | p < 0.05 |
|  |  | Chaiard, 2018[32] | Thailand | Cross-sectional | n=233 | Full-time nurses | OR = 4.25 | 1.74 to 10.38 | p < 0.001 |
| **Work over-commitment** | Excessive focus compromises safety awareness (**Ind**) | Wang, 2019[28] | China | Cross-sectional | n=1,956 | Hospital HCWs | r = 0.45 | 0.41 to 0.49□ | p < 0.01 |
|  |  | Bun, 2024[9] | France | Longitudinal | n=730 | Hospital HCWs | RR = 1.96 | 1.56 to 2.47 | p < 0.001 |
| **Nurse occupation** | Higher exposure to invasive procedures (**Ind**) | Jahic, 2018[33] | Bosnia-Herzegovina | Cross-sectional | n=1,031 | Hospital HCWs | F = 5.62 | NR | p = 0.004 |
|  |  | Bun, 2024[9] | France | Longitudinal | n=730 | Hospital HCWs | RR = 2.90 | 1.32 to 4.45 | p = 0.012 |
| **Younger age** | Inexperience and risk-taking behaviour (**Ind**) | Hassanipour, 2021[30] | Global | Meta-analysis | 43 articles (n=14,817) | Healthcare personnel | OR = 2.75 | 2.27 to 3.33 | p < 0.001 |
|  |  | Bun, 2024[9] | France | Longitudinal | n=730 | Hospital HCWs | RR = 4.25 | 1.20 to 9.94 | p = 0.026 |

Effect sizes: d, Cohen's d; IRR, incidence rate ratio; NR, not reported; OR, odds ratio; RR, relative risk; r, correlation coefficient; $\beta$ , standardised regression coefficient; $\gamma$ , gamma coefficient. Level: Ind, individual; Org, organisational; Ext, external. □ Calculated/transformed from reported data.

#### Psychological Mediators

**Stress-AEB relationship:** As shown in Table 2C, effect sizes ranged from OR = 1.12 (95% CI: 1.05-1.19) among Spanish HCWs[5] to r = 0.53 (95% CI: 0.50-0.56) in a Chinese cohort [28].

**Fatigue-AEB relationship:** Table 2C demonstrates stronger associations between fatigue and AEB:

- OR = 4.25 (95% CI: 1.74-10.38) among Thai nurses[32]
- IRR = 2.94 (95% CI: 1.71-5.07) in a North American case-crossover study[31]

#### Organisational Risk Factors

Multiple organisational factors showed substantial effects on AEB risk (Table 2C):

- **Irregular work schedules:** RR = 3.18 (95% CI: 1.83-5.11) in STRIPPS[9]; OR = 1.46 (95% CI: 1.15-1.86) in US nurses[29]
- **Rotating shifts:** RR = 3.11 (95% CI: 1.72-4.83) in STRIPPS[9]; OR = 2.16 (95% CI: 1.47-3.15) in global meta-analysis[30]
- **Use of external personnel:** RR = 2.02 (95% CI: 1.19-3.35)[9]

#### Individual Amplifying Factors

As detailed in Tables 2A and 2B:

- **Sickness presenteeism:** β = 4.03 (95% CI: 2.84-5.22) for association with fatigue; β = 1.61 (95% CI: 0.55-2.66) for association with stress[8].
- **Work overcommitment:** RR = 1.96 (95% CI: 1.56-2.47) for AEB risk[9]; r = 0.45 (95% CI: 0.41-0.49) with AEB in Chinese HCWs[28].

### Final DAG Model

Figure 2 presents the synthesised DAG illustrating causal pathways from organisational factors to AEB risk.

**Figure 2.**
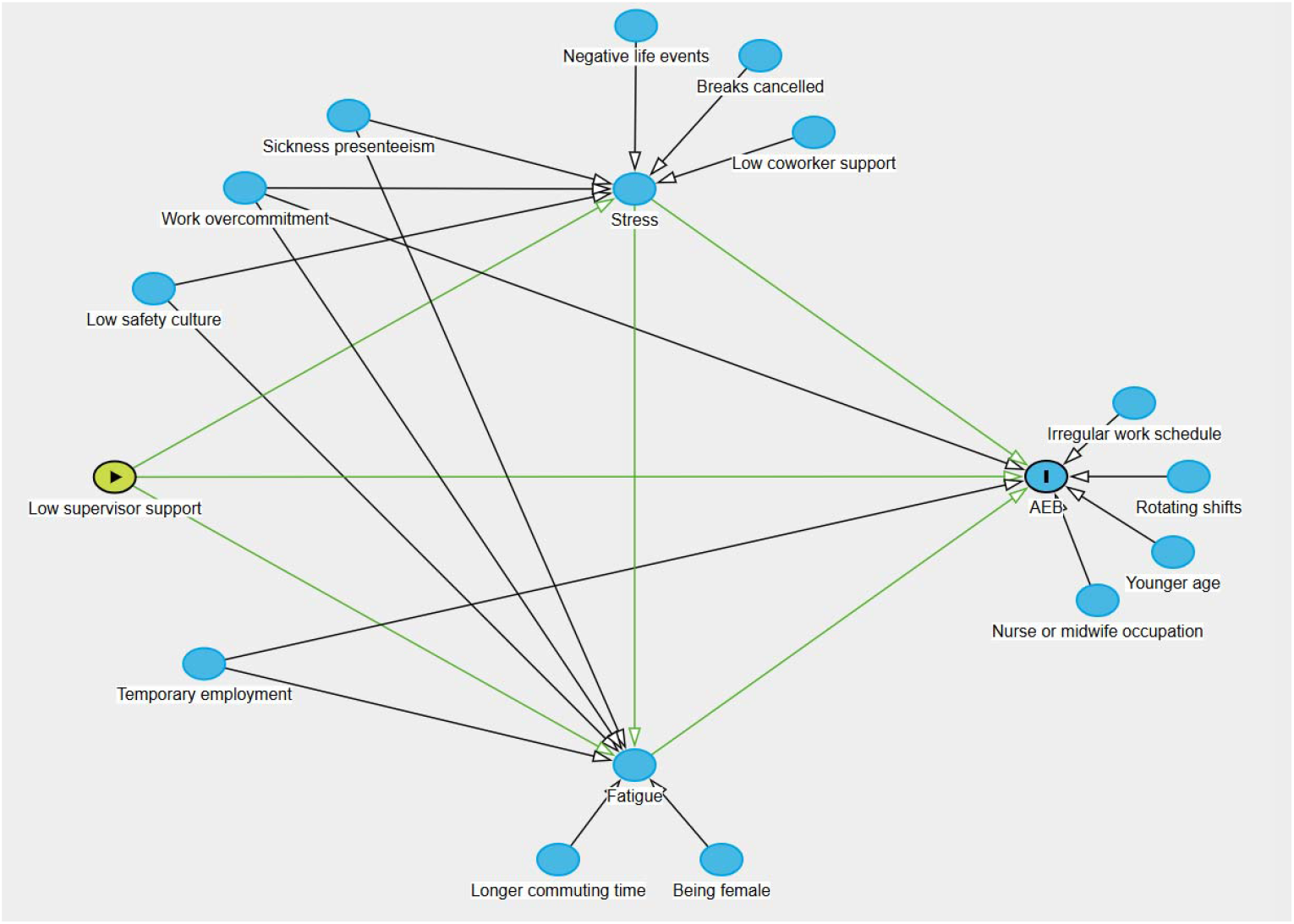
Directed Acyclic Graph showing causal pathways from supervisor support to Accidental Exposure to Blood

The DAG illustrates two primary pathways through which low supervisor support increases AEB risk:

1. **Direct pathway:** Supervisor support → AEB (RR = 1.16)
2. **Indirect pathways via psychological mediators:**
  ∘ Support → Stress → AEB
  ∘ Support → Fatigue → AEB

Additional factors identified in the model:

- **Organisational factors:** irregular schedules, rotating shifts, external personnel use
- **Individual factors:** presenteeism, overcommitment, age, occupation
- **Mediating variables:** stress and fatigue as central nodes connecting multiple pathways

## Discussion Principal Findings

A key finding is that the effect of low supervisor support is not merely direct but is predominantly mediated through the psychological pathways of stress and fatigue, two central occupational health outcomes.

Supervisor support thus plays a central role in HCW safety through both direct and indirect pathways to AEB risk. The identification of stress and fatigue as key mediators provides specific targets for intervention, while the quantification of effect sizes helps prioritise which relationships may yield the greatest impact if modified. The substantial variation in AEB incidence across specialties (2.1-7.1 per 100 visits) underscores the importance of context-specific approaches to prevention.

### Comparison with Existing Literature

Our findings align with previous research on organisational determinants of occupational safety. The observed relationship between supervisor support and psychological outcomes corroborates recent meta-analyses demonstrating the critical role of leadership in workplace safety [6]. The protective effect of supervisor support (RR=1.16 for AEB) is consistent with studies from diverse healthcare settings, though direct comparisons are limited by varying outcome measures and populations[28].

The stronger association of fatigue with AEB (OR ranging from 2.94 to 4.25) compared to stress (OR 1.12 to 1.53) aligns with recent study highlighting fatigue as a primary risk factor for occupational injuries[34]. This finding is particularly relevant given the increased focus on HCW fatigue following the COVID-19 pandemic, where extended shifts and staffing shortages have exacerbated baseline fatigue levels.

Our identification of sickness presenteeism as a major amplifying factor (β=4.03 for fatigue) resonates with emerging evidence on its detrimental effects. Studies have shown presenteeism rates ranging from 40-86% among HCWs, with significant associations with both burnout and reduced work ability[35]. The strong effect on fatigue suggests that working while ill creates a vicious cycle of exhaustion and increased injury risk.

### Theoretical Implications

From an occupational health perspective, our DAG supports the Job Demands-Resources (JD-R) model, demonstrating how supervisor support functions as a critical resource buffering job demands[36]. The parallel pathways through stress and fatigue suggest that organisational stressors affect multiple physiological and psychological systems simultaneously, consistent with occupational health models that emphasise the multifactorial nature of workplace injuries[37].

The complex interaction between organisational and individual factors illustrated in our DAG challenges simple linear models of occupational safety. Instead, it supports a systems approach recognising multiple, interconnected determinants operating at different levels[37]. This complexity underscores why single-intervention approaches often fail to substantially reduce AEB rates.

### Practical Implications

Our findings provide occupational health practitioners and prevention teams with clear target for intervention. First, investing in supervisor training programs that emphasise supportive leadership behaviours could yield multiple benefits. Previously, leadership interventions have shown 20-30% reductions in employee stress and improvements in safety outcomes[38]. Given the relatively modest direct effect (RR=1.16), interventions should target multiple pathways simultaneously.

Second, addressing presenteeism requires cultural change that should be guided by occupational health teams. Organisations should examine factors driving presenteeism, including staffing levels, sick leave policies, and implicit expectations about working while ill[35]. The strong association with fatigue suggests that presenteeism policies could be particularly important for fatigue management.

Third, the substantial effects of scheduling factors (RR>3.0 for irregular schedules and rotating shifts) indicate that roster optimization, a key lever for occupational physicians, could significantly reduce AEB risk. Innovations in predictive scheduling and fatigue risk management systems offer promising approaches[39].

### Strengths and Limitations

Strengths include the multi-site design, comprehensive data collection, and systematic approach to DAG construction combining empirical data with literature synthesis. The use of previously published multivariate-adjusted effect sizes strengthens causal inference by accounting for measured confounders.

Several limitations must be acknowledged. First, while our DAG respects temporal ordering where possible, some relationships rely on cross-sectional data, limiting causal inference. Additionally, effect sizes from different studies used varying metrics (OR, RR, β, r), requiring careful interpretation when comparing magnitudes across pathways.

Second, AEB self-reporting likely underestimates true incidence. Studies suggest only 40-50% of AEB are reported through official channels[14]. However, self-reported data may better capture events HCWs perceive as significant, regardless of formal reporting.

Third, unmeasured confounders may influence observed relationships. We lacked data on specific safety training, personal protective equipment availability, and individual risk-taking behaviours. The ecological correlation between ward-level supervisor support and AEB rates did not reach statistical significance, possibly reflecting limited statistical power with only 32 ward-level observations.

Regarding generalisability, while our cohort was limited to four Parisian university hospitals, the consistency of our findings with international literature suggests broader applicability. The identified pathways involving support, stress, and fatigue may be relevant for other occupational injuries such as musculoskeletal disorders in high-risk sectors. This suggests that the underlying causal model could have broader applicability in occupational safety research.

### Future Research Directions

Longitudinal studies with repeated measures could better establish temporal relationships and test whether changes in supervisor support lead to predicted changes in downstream outcomes. Intervention studies targeting supervisor training with concurrent measurement of stress, fatigue, and AEB would provide stronger causal evidence.

Additionally, exploration of potential moderators – such as ward type, baseline safety culture, or individual resilience factors – could identify contexts where interventions might be most effective. Economic evaluations of multi-level interventions would help healthcare organisations prioritise resource allocation.

The relationships identified in our DAG may extend to other safety outcomes. Future research should examine whether similar pathways influence hand hygiene compliance, medication errors, or patient safety incidents[40].

## Conclusions

This study provides a comprehensive occupational health causal model linking organisational factors to AEB risk in healthcare settings. Low supervisor support emerges as a critical upstream factor influencing occupational safety through stress and fatigue pathways. The convergence of multiple risk factors highlights the need for multilevel interventions that should be implemented in collaboration with occupational services.

Healthcare organisations should prioritise supportive supervision, fatigue management, and systematic approaches to scheduling as integral components of comprehensive occupational safety and health programs. By addressing root organisational causes rather than focusing solely on individual behaviours, healthcare systems can create safer environments for their workforce.

## Data Availability

All data produced in the present study are available upon reasonable request to the authors

## Acknowledgements

We would like to acknowledge the support and funding provided by Assistance Publique – Hôpitaux de Paris (Département de la Recherche Clinique et du Développement). We would also like to express our gratitude to all the healthcare workers who participated in this study.

